# Predicting seizure outcome after epilepsy surgery: do we need more complex models, larger samples, or better data?

**DOI:** 10.1101/2023.02.13.23285866

**Authors:** Maria H Eriksson, Mathilde Ripart, Rory J Piper, Friederike Moeller, Krishna B Das, Christin Eltze, Gerald Cooray, John Booth, Kirstie J Whitaker, Aswin Chari, Patricia Martin Sanfilippo, Ana Perez Caballero, Lara Menzies, Amy McTague, Martin M Tisdall, J Helen Cross, Torsten Baldeweg, Sophie Adler, Konrad Wagstyl

## Abstract

**Objective:** The accurate prediction of seizure freedom after epilepsy surgery remains challenging. We investigated if 1) training more complex models, 2) recruiting larger sample sizes, or 3) using data-driven selection of clinical predictors would improve our ability to predict post-operative seizure outcome. We also conducted the first external validation of a machine learning model trained to predict post-operative seizure outcome.

**Methods:** We performed a retrospective cohort study of 797 children who had undergone resective or disconnective epilepsy surgery at a single tertiary center. We extracted patient information from medical records and trained three models – a logistic regression, a multilayer perceptron, and an XGBoost model – to predict one-year post-operative seizure outcome on our dataset. We evaluated the performance of a recently published XGBoost model on the same patients. We further investigated the impact of sample size on model performance, using learning curve analysis to estimate performance at samples up to *N*=2,000. Finally, we examined the impact of predictor selection on model performance.

**Results:** Our logistic regression achieved an accuracy of 72% (95% CI=68-75%, AUC=0.72), while our multilayer perceptron and XGBoost both achieved accuracies of 71% (95% CI_MLP_=67-74%, AUC_MLP_=0.70; 95% CI_XGBoost own_=68-75%, AUC_XGBoost own_=0.70). There was no significant difference in performance between our three models (all *P*>0.4) and they all performed better than the external XGBoost, which achieved an accuracy of 63% (95% CI=59-67%, AUC=0.62; *P*_LR_=0.005, *P*_MLP_=0.01, *P*_XGBoost own_=0.01) on our data. All models showed improved performance with increasing sample size, with limited improvements above *N*=400. The best model performance was achieved with data-driven feature selection.

**Significance:** We show that neither the deployment of complex machine learning models nor the assembly of thousands of patients alone is likely to generate significant improvements in our ability to predict post-operative seizure freedom. We instead propose that improved feature selection alongside collaboration, data standardization, and model sharing is required to advance the field.

## Introduction

Despite careful evaluation, up to one third of patients with drug-resistant epilepsy are not rendered seizure-free through surgery^1^. This underscores the need to identify which patients are likely to benefit from surgery before the intervention has been carried out.

Surgical candidate selection is typically decided by a multidisciplinary team. This form of expert clinical judgement relies on experience and available evidence, and achieves a moderate degree of accuracy when predicting surgical success.^2^ To aid clinical judgement, some studies have reported average estimates of seizure freedom for specific types of epilepsy (e.g. temporal lobe epilepsy).^1^ Other studies have focused on identifying multiple predictors of post-operative seizure outcome, without taking into account how these predictors may interact.^1^

In an effort to synthesize patient characteristics and provide objective predictions of seizure freedom, researchers have developed statistical models and calculated risk scores that can generate individualized predictions of outcome.^3–5^ These have included the Epilepsy Surgery Nomogram^3^, the modified Seizure Freedom Score^4^, and the Epilepsy Surgery Grading Scale.^5^ These tools do not, however, perform better than clinical judgment.^2,6^ Researchers are therefore increasingly turning to machine learning in an attempt to improve prediction accuracy.

Machine learning is being leveraged within the realm of clinical research at an exponential pace. The epilepsy surgery pathway generates a plethora of diverse data. As such, it would seem to create an ideal opportunity for the application of machine learning technology. Several machine learning models have indeed been developed to date to predict seizure outcome (**Table 1**). The majority of these models have, however, been trained on relatively small sample sizes (*N* < 100)^7–18^ and therefore have a high risk of ‘overfitting’ (a model overfits when it models the training dataset too closely, performing well on this dataset but consequently underperforming on new, ‘unseen’ datasets).^19,20^ Model training sets have also been comprised almost exclusively of temporal lobe surgery patients^7,8,10–13,15,17,18,21,22^, often relied on post-surgical factors^11,12,14,23^, and frequently utilized post-processing neuroimaging analyses that cannot be readily replicated by others.^7,9–11,13,16–18,21^ As such, many existing models may be difficult to incorporate into routine pre-surgical evaluation. Perhaps more importantly still, none of these models have been externally validated. It is therefore unknown how well they would perform if used by another surgery center, and whether their adoption as a replacement for traditional statistical modelling approaches is justified.

To advance this field, we asked whether 1) more complex models, 2) larger sample sizes, or 3) better selection of clinical predictors would improve our ability to predict post-operative seizure outcome (**Fig. 1**). To address the first question, we trained three different models, a traditional logistic regression and two machine learning models, to predict seizure outcome on our dataset. We also tested the performance of an external, pre-trained machine learning model^23^ on our dataset, and compared its performance to that of our models. To address the influence of sample size, we investigated how varying sample size – both within and extrapolating beyond our current cohort – impacted model performance. To address the influence of number and type of clinical predictors, we investigated how the inclusion of different predictors affected model performance.

**Figure 1.**
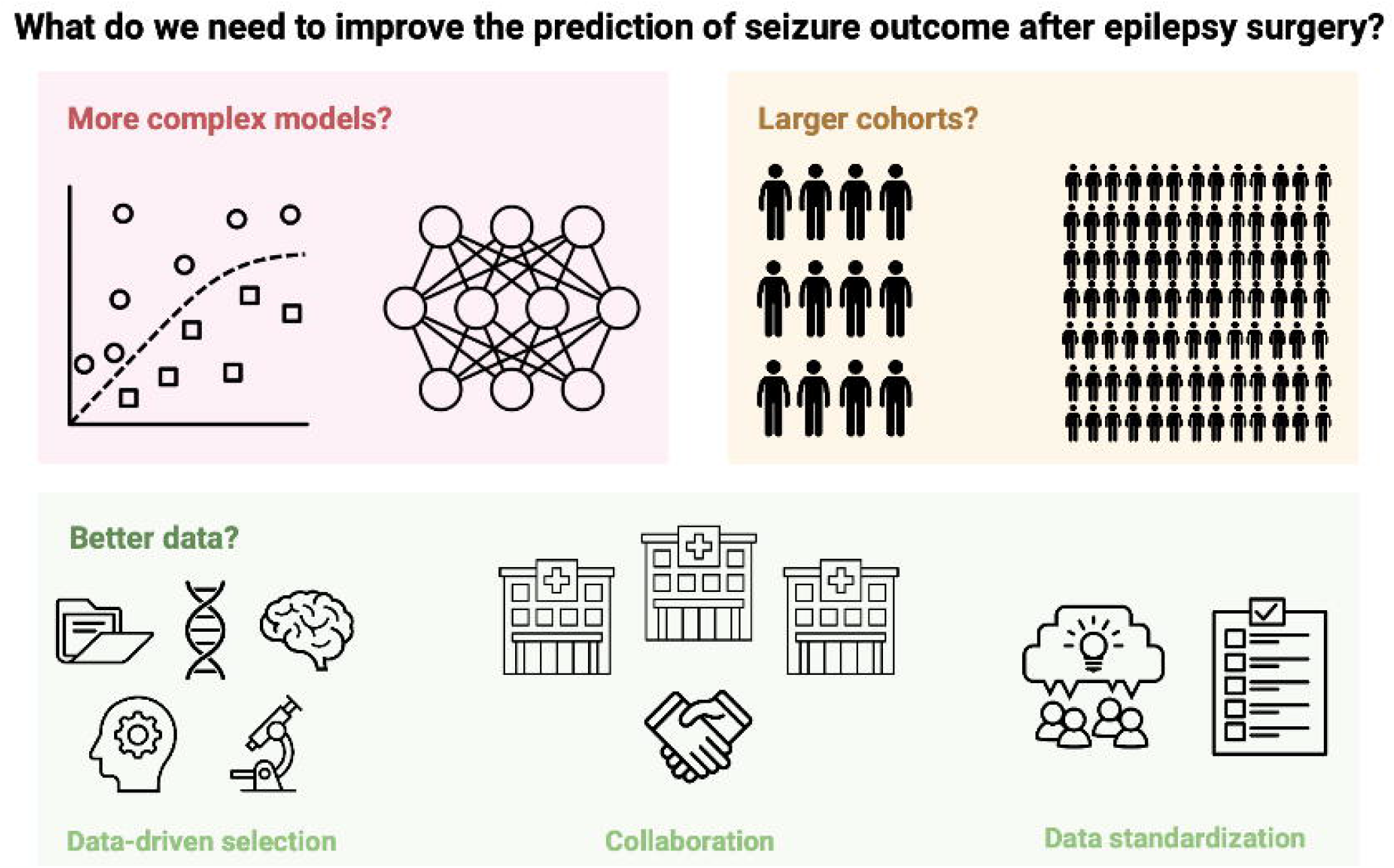
Study overview. We investigated the impact of model type, sample size, and feature selection on our ability to accurately predict post-operative seizure outcome.

## Materials and methods

### Patient cohort

We retrospectively reviewed medical records for all children who underwent epilepsy surgery at Great Ormond Street Hospital (GOSH; London, UK) from 2000 through 2018. We included patients who underwent surgical resection or disconnection. We excluded palliative procedures (corpus callosotomy and multiple subpial transections), as well as neuromodulation (deep brain stimulation and vagal nerve stimulation) and thermocoagulation procedures. If patients had undergone multiple surgeries over the course of the study period, we included only their first surgery.

### Dataset description

We retrieved medical records and extracted the following information: patient demographics, epilepsy characteristics, pre-operative MRI findings, pre-operative interictal and ictal EEG characteristics, pre-operative antiseizure medication (ASM; including both total number of ASMs trialed from time of epilepsy onset to time of pre-surgical evaluation, as well as number of ASMs at time of pre-operative evaluation), surgery details, genetic results, and histopathology diagnosis. A complete list of variables extracted and information about how we categorized these data can be found in **Supplementary Material** (p. 2-6).

We classified patients as either seizure-free (including no auras) or not seizure-free at one-year post-operative follow-up. We also recorded if patients were on, weaning or off ASMs at this time-point.

### Statistical analysis

We calculated the descriptive statistics for the cohort and presented these using mean with standard deviation, median with interquartile range, and count with proportion, as appropriate.

We checked if continuous data were normally distributed using Shapiro-Wilk tests.^24^ None of the continuous variables were normally distributed. We therefore investigated associations between demographic, clinical and surgical variables using Mann-Whitney U, Kruskal-Wallis H, Chi-square test of independence, and Spearman’s rank correlation coefficient, as appropriate. All tests were two-tailed and we set the threshold for significance a priori at *P* ≤ 0.05. We corrected for multiple comparisons using the Holm method.^25^

We performed univariable logistic regression analyses to investigate which clinical variables predicted seizure outcome at one-year post-operative follow-up. In the case of categorical variables, the group known to have the highest seizure freedom rate (according to past literature) was used as the reference category. All other groups were then compared to this reference category to determine if they were significantly less (or more) likely to achieve seizure freedom through surgery. For example, ‘unilateral MRI abnormalities’ was selected as the reference category for the categorical variable ‘MRI bilaterality’, and we investigated whether those with ‘bilateral MRI abnormalities’ were significantly less (or more) likely to be seizure-free after surgery. We again corrected for multiple comparisons using the Holm method.^25^

#### Effect of model type on model performance

We performed a multivariable logistic regression (LR) with independent variables that 1) could be obtained pre-surgically and 2) were found to be predictive of seizure outcome. We developed a second version of this model, in which MRI diagnosis was replaced with histopathology diagnosis, to determine if this affected model accuracy.

We used the same predictors to train two machine learning models: a multilayer perceptron (MLP) model and an XGBoost model. We chose an MLP due to its high predictive performance, allowing for non-linear interactions between input variables. We trained the MLP with two hidden layers and ten hidden neurons, respectively, balancing the need for sufficient complexity to learn feature interactions across multiple features, while limiting the capacity of the network to overfit to the training data. We chose an XGBoost model to ensure that we could compare the performance of this to the performance of the XGBoost model published by Yossofzai *et al*.^23^

After training our own three models, we applied the XGBoost model by Yossofzai *et al*.^23^ to the same patient cohort. We evaluated the performance of all models using stratified 10-fold cross-validation. We used a stratified approach to address the outcome imbalance observed in our cohort. We calculated the null accuracy (the accuracy the model would achieve if it always predicted the more commonly occurring outcome in our cohort, i.e. seizure-free), the tested model accuracy, and the area under the ROC (Receiver Operating Characteristic) curve (AUC) for each model. We reported both the mean AUC obtained across all 10 folds as well as the AUC obtained from each individual fold. We compared the accuracies of the respective models using McNemar’s test.

#### Effect of sample size on model performance

We investigated how sample size affected model performance by using a previously described learning curve analysis approach.^26^ First, we trained our models on ten different sample sizes, starting at *N*=70 and finishing at *N*=700 patients. At each sample size, we evaluated model performance, specifically model accuracy. This allowed us to create a learning curve, plotting model performance against sample size. We then chose an inverse power law function to model the learning curve. We used this function to predict model performance on expanded sample sizes of up to *N*=2,000.

#### Effect of clinical predictors on model performance

We explored how the number of included predictors, as well as their nature, affected model performance. We used the coefficients from our univariable logistic regression analyses to determine how informative different predictors were. We then added significant predictors one-by-one into our models, from the most informative to the least informative. At each point, we plotted model AUC and confidence intervals (obtained across the 10 folds).

We performed all statistical analyses and visualizations in Python version 3.7.2 and R version 3.6.3. Our MLP and XGBoost models were implemented using the scikit-learn library.^27^

## Results

### Patient cohort

A total of 797 children were identified as having undergone first-time surgical resection or disconnection. Demographic information and clinical characteristics for these patients are displayed in **Supplementary Table 1**. Data relating to semiology (past seizures and seizures at time of pre-surgical evaluation) as well as interictal and ictal EEG characteristics are displayed in **Supplementary Table 2**. Genetic diagnoses are listed in **Supplementary Tables 3** and **4**.

Seizure outcome at one-year follow-up was available for 709 patients, of which 67% were seizure-free. Of these, 51% were on ASM, 34% were weaning ASM, and 15% were on no ASM.

### Relationships between variables

Relationships between demographic, clinical and surgical variables are displayed in **Fig. 2**. Full statistics are reported in **Supplementary Table 5**.

**Figure 2.**
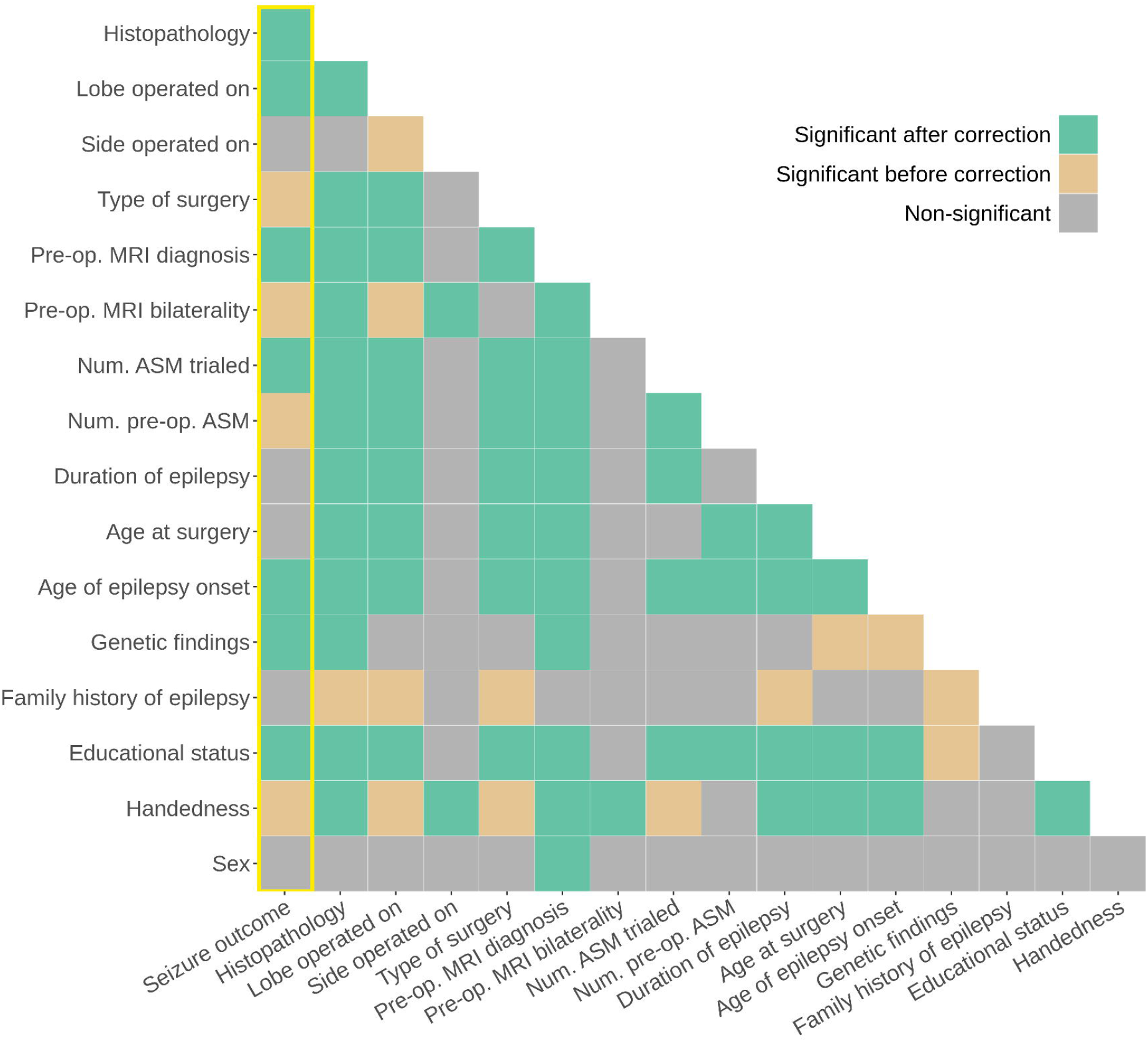
Relationships between demographic, clinical and surgical variables. Relationships are shown both before and after correction for multiple comparison using the Holm method. We have highlighted relationships with seizure outcome using a yellow box.

### Univariable logistic regression analyses

Univariable logistic regression analyses identified the following features as predictive of one-year post-operative seizure freedom: handedness, educational status, genetic findings, age of epilepsy onset, history of infantile spasms, spasms at time of pre-operative evaluation, number of seizure types at time of pre-operative evaluation, total number of ASMs trialed (from time of epilepsy onset to time of pre-operative evaluation), MRI bilaterality (unilateral versus bilateral abnormalities), MRI diagnosis, type of surgery performed, lobe operated on, and histopathology diagnosis (**Supplementary Table 6**).

### Effect of model type on model performance

#### Logistic regression models

Our multivariable LR achieved an accuracy of 72% (95% CI=68-75%) and an AUC of 0.72 (range across the 10 folds: 0.64-0.82). When we assessed whether substituting MRI diagnosis with histopathology diagnosis would improve model performance, we found that this alternative LR achieved a similar accuracy of 73% (95% CI=69-79%; AUC=0.72; range across the 10 folds: 0.60-0.77). There was no significant difference in performance between the LR that included MRI diagnosis and the LR that included histopathology diagnosis (McNemar’s test, chi-square=0.1, *P*=0.8). This was likely due to the high degree of overlap between MRI and histopathology diagnosis (**Supplementary Fig. 1**).

#### Multilayer perceptron and XGBoost models

Our MLP achieved an accuracy of 71% (95% CI=67-74%) and an AUC of 0.70 (range across the 10 folds: 0.63-0.82). Our XGBoost also achieved an accuracy of 71% (95% CI=68-75%) and an AUC of 0.70 (range across the 10 folds: 0.62-0.83).

#### External XGBoost model

When we applied the XGBoost model developed by Yossofzai *et al*.^23^ to our data, it achieved an accuracy of 63% (95% CI=59-67%) and an AUC of 0.62.

#### Comparison of model performances

The AUCs of the respective models are compared in **Fig. 3A**. There was no significant difference in performance between our LR and MLP (McNemar’s test, chi-square=0.8, *P*=0.4), our LR and XGBoost (McNemar’s test, chi-square=0.1, *P*=0.8), or our MLP and XGBoost (McNemar’s test, chi-square=0.1, *P*=0.8).

**Figure 3.**
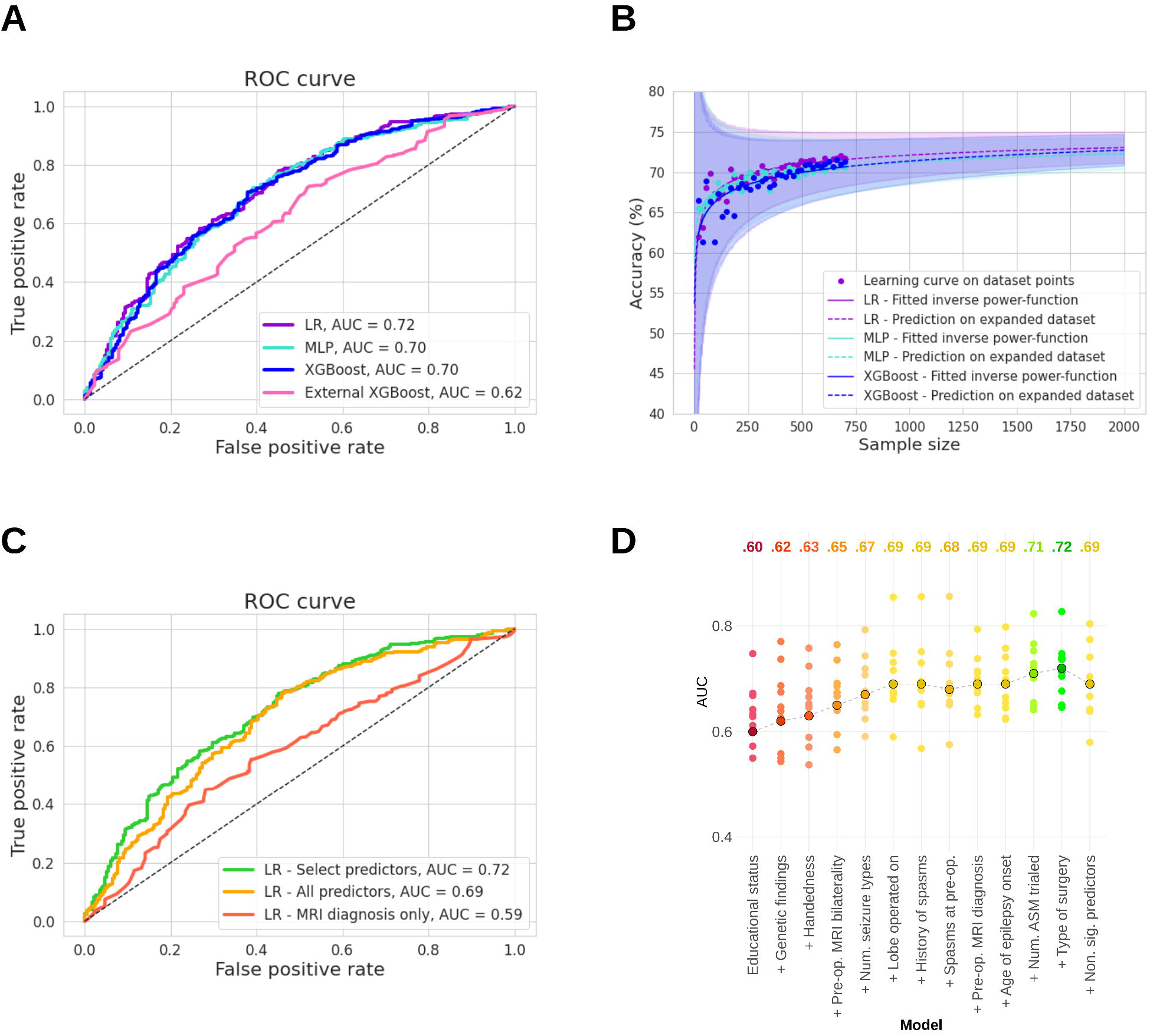
Impact of model type, sample size, and selection of clinical variables on model performance. **(A)** Receiver operating characteristic (ROC) curves showing model performances. There was no significant difference in performance between our LR (purple), MLP (turquoise), and XGBoost (blue) models. All of our models performed significantly better than the XGBoost model recently developed by Yossofzai *et al*.^23^ (pink). **(B)** Effect of sample size on model performance (accuracy). There was an improvement in model performance with increasing sample size for our LR, MPL and XGBoost models, up until around *N*=400. After this point, the model showed only marginal gains in performance. Extrapolating performance for sample sizes up to *N*=2,000 did not predict substantial improvement in model performance for any of our models. **(C)** Receiver operating characteristic (ROC) curves showing model performance for our LR models containing 1) only MRI diagnosis (red), 2) all predictors (orange), and 3) predictors identified through data-driven feature selection (green). Data-driven selection involved including only predictors that were significantly predictive of one-year seizure outcome as identified in univariable logistic regression analyses. Corresponding ROC curves showing model performances for our MLP and XGBoost models are displayed in **Supplementary Fig. 2** and **3. (D)** Effect of data-driven feature selection on model performance (AUC). Variables found to be significantly predictive of seizure outcome from univariable logistic regression analyses were added one-by-one to the LR, from most information to least informative according to their coefficients. Model performance increased in line with the predictors being added. Adding the remaining predictors collected for the study, i.e. those that were not significantly predictive of seizure outcome, worsened model performance (far right). Points circled in black represent mean AUC obtained across all 10 folds. Non-circled points represent the AUCs obtained from each of the individual 10 folds.

All three models performed better than the external XGBoost model (McNemar’s test_LR_, chi-square=8.0, *P*=0.005; McNemar’s test_MLP_, chi-square=6.4, *P*=0.01; McNemar’s test_XGB own_, chi-square=6.8, *P*=0.01). Our LR, MLP and XGBoost models also performed significantly better than model null accuracy (McNemar’s test_LR_, chi-square=8.7, *P*=0.003; McNemar’s test_MLP_, chi-square=5.3, *P*=0.02; McNemar’s test_XGB own_, chi-square=7.6, *P*=0.006), whereas the external XGBoost model did not (McNemar’s test_XGB external_, chi-square=0.6, *P*=0.4).

### Effect of sample size on model performance

Increasing our sample size within the limits of our cohort improved the performances of all our models (**Fig. 3B**). This was, however, only true up until around *N*=400, at which point performance started to plateau for all models. Expanding our cohort beyond its current size, up to *N*=2,000, did not substantially improve the performances for any of our models (**Fig. 3B**).

### Effect of data inclusion on model performance

We found that adding more predictor features improved the performances of all models (**Fig. 3C** and **Supplementary Fig. 2** and **3**). However, the greatest accuracy was achieved when data-driven feature selection was used to filter which clinical predictors should be included in the models (i.e. when the models included only the variables that were found to be significantly predictive of seizure outcome in our univariable logistic regression analyses; **Fig. 3D**). When we added variables that were not significantly predictive of seizure outcome in our univariable logistic regression analyses, model performance worsened (**Fig. 3D**).

## Discussion

Up to one third of patients do not achieve seizure freedom through epilepsy surgery despite careful evaluation.^1^ There has been a long-standing history of trying to identify these patients pre-operatively, both through traditional statistical modelling approaches and more complex machine learning techniques (**Table 1**). These attempts have, however, had limited success. In this study, we explored if we could improve our ability to predict seizure outcome by training more complex models, recruiting larger training sample sizes, or incorporating more or different types of clinical predictors.

To investigate the effect of model type on our ability to predict seizure outcome, we trained three different models, a logistic regression (LR) and two machine learning models – a multilayer perceptron (MLP) and an XGBoost – on the same cohort. We showed that our LR performed as well as our MLP and XGBoost models. Importantly, we also applied a recently published XGBoost model by Yossofzai *et al*.^23^ to our cohort, and found that this model performed worse than our models (AUC=0.62 versus AUC=0.70-0.72). It also performed worse on our cohort compared to the cohorts it was trained and tested on (AUC=0.62 versus AUC=0.73-0.74).

To address the value of larger patient sample sizes, we investigated model performance on a range of sample sizes, up to *N*=2,000. We found that the performances of all models improved until around *N*=400, after which point they began to plateau.

To address the influence of clinical predictors, we varied both the number of predictors included in the models as well as the nature of these predictors. We demonstrated that using data-driven feature selection (i.e. including only variables that were predictive of seizure outcome in univariable logistic regression analyses) resulted in the best model performance, while including all collected predictors led to a deterioration in model performance. Interestingly, neither EEG nor semiology characteristics were predictive of seizure outcome in our univariable logistic regression analyses and therefore not included in our models.

### The illusory superiority of more complex models

There is a growing tendency to favor machine learning technology over traditional statistical modelling approaches when training models to predict post-operative seizure outcome. This is presumably due to an assumed superiority of highly sophisticated or complex models. As a result, a plethora of machine learning techniques have been deployed (**Table 1**). It is, however, also increasingly recognized that the potential gains in predictive accuracy that have been attributed to more complex algorithms may have been inflated^20,28^, and that minor improvements observed “in the laboratory” may not translate into the real-world.^20^

Previous studies that have used both machine learning techniques and traditional statistical modelling approaches to predict post-operative seizure outcome have found that logistic regression models perform as well as, or even better than, machine learning ones.^11,15,22^ To our knowledge, only one study by Yossofzai *et al*.^23^ has found that a machine learning model outperforms a logistic regression; however, this was a 0.1-0.2 difference in AUC (0.72 versus 0.73 in the train dataset; 0.72 versus 0.74 in the test dataset). This small improvement is unlikely to deliver an advantage in clinical practice. At the same time, using machine learning models introduces complexity, which in turn complicates their interpretation, implementation and validation, and increases the risk of overfitting.

### Larger samples mean higher accuracy… but only up until a certain point

There exists a general consensus in the machine learning community that more data, or larger sample sizes, equates to better model performance.^29,30^ However, researchers have started to show that this is not always the case.^31^ We found that expanding our cohort beyond its current size (*N*=797) nearly three-fold did not provide meaningful gains.

Estimating the point of diminishing returns is invaluable because – whilst there is an abundance of unlabeled clinical data in our era of Big Data – (human) annotated clinical data remains scarce. Its creation is time-consuming and requires the expertise of several clinical groups. Nevertheless, annotated datasets are essential in the creation of (supervised) learning algorithms. Generating learning curves can therefore inform researchers of the relative costs and benefits of adding additional annotated data to their model.^32^ Still, it is important to note that this learning curve is only an estimate and that actual model performance could exceed these predictions.

### In pursuit of (geographical) model generalizability

Machine learning in clinical research is placing an increasing emphasis on model generalizability, where the highest level of evidence is achieved from applying models externally – to new centers. When we tested the model by Yossofzai *et al*.^23^ on our data, we found that it did not generalize well. This may at first glance seem surprising, as there is a striking similarity between our cohort and the cohort of Yossofzai *et al*.^23^ – not only in terms of sample size, but also in terms of patient characteristics and variables found to be predictive of outcome. However, it also highlights a common issue related to the use of machine learning, namely the tendency for models to overfit to local data. We therefore expect that a similar decrease in model performance would be demonstrated if another center were to use the machine learning models that we trained.

Different epilepsy surgery centers show variation in which diagnostic and therapeutic procedures are available, for which patients they are requested, and with which specifications they are carried out.^33^ Local practices also influence how data are annotated. Clinical data are interpreted by experts who assign a wide range of labels, from MRI diagnosis to epilepsy syndromes. Whilst official classification systems for annotation procedures exist^34^–^39^, individual studies often choose to – or are forced to – categorize their data ad hoc, often due to the restraints introduced by the retrospective nature of their data. Furthermore, not all experts will agree on the same label, which is evidenced by a lack of agreement regarding interpretation of EEG^40^–^42^, MRI^43^, PET^43^ and histopathological data^34^. It is thus possible that while our cohort and the cohort of Yossofzai *et al*.^23^ look similar on the surface, they may represent patients who have been characterized in a subtly different manner.

### Limitations of the current study

The primary limitation of our study is that it is a retrospective study, which uses data originally obtained to understand patient disease and support clinical care, rather than to enable data analysis. These data are therefore at risk of being biased and incomplete.

#### Biased data

Presurgical evaluation is largely standardized in that all patients undergo a full clinical history, structural MRI, and scalp- or video-EEG, but the extent of further investigations will be patient dependent.^44^ To mitigate the occurrence of bias, we used a minimal dataset, which included only clinical variables typically obtained for all epilepsy surgery patients. As such, we did not train our model using positron emission tomography (PET), single-photon emission computed tomography (SPECT), magnetoencephalography (MEG), or functional MRI (fMRI) measures. One exception to this was the inclusion of genetic diagnosis, which we included despite not all patients having undergone genetic testing. The predictive value of genetic information in surgery candidate selection has not been systematically investigated.^45^ We therefore sought to contribute to this emerging area of research and provide initial evidence for its importance.

#### Incomplete data

Related to the limitation of biased data is the limitation of incomplete data. Similar to past retrospective studies that have developed models for the prediction of seizure outcome after epilepsy surgery, we had a considerable amount of missing data. There are multiple ways of handling incomplete datasets, including deleting instances or replacing them with estimated values – a method known as imputation. Imputation techniques must, however, be used with caution, as they have limitations and can impact model performance.^46^ We therefore chose to drop instances where continuous data points were missing before including them into the model training datasets, and classified missing categorical data points as such, rather than using imputation.

### Moving forward

Taken together, our findings suggest that 1) traditional statistical approaches such as logistic regression are likely to perform as well as more complex machine learning models (when using clinical predictors similar to those described here) and have advantages in interpretability, implementation and generalizability; 2) collecting a large sample is important because it improves model performance and reduces overfitting, but including more than a thousand patients is unlikely to generate significant returns on datasets similar to ours; 3) model improvement is likely to come from data-driven feature selection and exploring the inclusion of features that have thus far been overlooked or not undergone external validation due to barriers in study replication (discussed below).

Based on these findings, we make recommendations to advance our ability to predict seizure outcome after epilepsy surgery (**Table 2**). Surgery centers around the world must collaborate to produce high-quality data for *research* purposes. Although models trained on single center data are likely to produce higher model performances than multicenter datasets, they may not be suitable for use by other surgery centers. Critically, data must be collected and curated in a standardized manner, as highlighted by experts^47^ and similar to recent multicenter endeavours.^22,48,49^ Here, it will be important to distinguish between investigating variables that may be predictive of outcome and identifying variables that can (feasibly) be included as predictors in a clinical decision-making tool. For the purpose of developing a clinical decision-making tool, we suggest including only variables that are routinely collected for all epilepsy surgery patients at most centers, to avoid introducing bias into the model. In other words, researchers should carefully consider the added value of modalities such as MEG, PET, SPECT and fMRI. Importantly, only variables obtained prior to surgery should be included in the model, as the aim is to create a predictive model. This means excluding variables such as post-operative measurement of resection and histopathology diagnosis. Reassuringly, we have shown that MRI diagnosis provides similar information to histopathology diagnosis. We also echo past recommendations^45^ in that we suggest avoiding variables that have repeatedly failed to predict outcome, as these have been shown to worsen model performance.

It is unlikely that clinical information alone will procedure high model performance, as demonstrated both here and by previous studies (**Table 1**). Instead, better data must also entail new data. The inclusion of additional predictors to improve model performance may involve extracting quantitative features from pre-operative MRI or EEG (as several studies detailed in **Table 1** have done), characterizing the epileptogenic network through computational modelling^50^, measuring lesion overlap with eloquent cortex^51^, or adopting a network analysis approach.^52^

It is important that all model software is made available – either as ready-to-use tools or openly shared code on platforms such as GitHub. Past studies have reported models capable of achieving accuracies as high as 90-100% using features extracted from MRI and EEG (**Table 1**); however, none of these findings can be reproduced, nor can any of these models be adopted by other centers, as there is insufficient information about how they were generated. Yossofzai *et al*.^23^ are to be commended for sharing their model in a way that allowed for it to be externally tested by ourselves and others.

## Conclusions

Accurate prediction of seizure outcome after epilepsy surgery remains difficult. We highlight the importance of comparing traditional statistical modelling to complex machine learning techniques, as we show that these two approaches may perform equally well. We also demonstrate the importance of performing external validation of machine learning models, as we show that algorithms may underperform on other centers’ data. Based on our findings, we present recommendations for future research, including the need for epilepsy services to collaborate in the creation of standardized datasets, the value of carefully choosing predictor variables for modelling, and the benefit of sharing models and code openly.

## Data Availability

The study's data dictionary, statistical analysis plan, and analytic code will be made available on GitHub (https://github.com/MariaEriksson/Predicting-seizure-outcome-paper). The full data are not publicly available due to their sensitive nature. Deidentified data will be made available from the corresponding author upon reasonable request.

## Glossary

ASM: antiseizure medication
AUC: Area under the (ROC) curve
CI: confidence interval
CNV: copy number variation
EEG: Electroencephalography
fMRI: Functional magnetic resonance imaging
IQR: interquartile range
LR: Logistic regression
MEG: Magnetoencephalography
MLP: Multilayer perceptron
MRI: Magnetic resonance imaging
OR: odds ratio
PET: Positron emission tomography
ROC: Receiver operating characteristic
SNV: single nucleotide variation
SPECT: Single-photon emission computed tomography

